# Neonatal mortality in The Gambia and Burkina Faso: insights to incidence and risk factors from clinical trial data

**DOI:** 10.1101/2025.11.02.25339325

**Authors:** Usman N Nakakana, Toussaint Rouamba, Bully Camara, Joel D. Bognini, Nathalie Beloum, Athanase M. Somé, Fatoumata Sillah, Guétawendé J W Nassa, Madikoi Danso, Edmond Yabré Sawadogo, Joquina C. Jones, Shashu Graves, Diagniagou Lankoandé, Ebrahim Ndure, Yusupha Njie, Christian Bottomley, Umberto D’Alessandro, Halidou Tinto, Helen Brotherton, Anna Roca

**Author notes:** Joint first authors (Usman N Nakakana, Toussaint Rouamba). Corresponding author & contact details: Dr Helen Brotherton.

## Abstract

**Introduction:** Neonatal mortality remains unacceptably high globally, particularly in Sub Saharan Africa. We evaluated the incidence and risk factors for neonatal mortality in The Gambia and Burkina Faso to provide context specific evidence to inform public health policy and programmes in West Africa.

**Methods:** We conducted secondary data analyses of the PregnAnZI2 trial which was a phase III, double blind, randomised clinical trial implemented across 10 government health facilities in urban Gambia and rural Burkina Faso. Pregnant women without known acute or chronic health conditions were enrolled during active labour and their neonates were actively followed up to 28 postnatal days. We calculated Neonatal Mortality Rates (NMR) and generated Kaplan-Meier survival curves to assess the timing of neonatal deaths. Logistic regression was applied to identify risk factors associated with early (0 to 72 hours) neonatal deaths, adjusted for potential confounders.

**Results:** 12,105 neonates born from 11,983 women were included in the analyses. Overall, 144 infants died during the neonatal period (NMR 11.5 per 1,000 live births), with 82% (118/144) of deaths occurring during the first 72 hours (early neonatal deaths). Risk factors for early neonatal mortality were low Apgar score at 1 minute (aOR 49.85, 95% CI 32.2 to 77.6) and 5 minutes (aOR 92.38, 95% CI 56.6 to 151.3) which are proxy for intrapartum related asphyxia, congenital malformations (aOR 11.44, 95% CI 6.4 to 19.4), and low birth weight (aOR 4.09, 95% CI 2.6 to 6.3). Other risk factors associated with early neonatal mortality included maternal history of stillbirth (cOR 4.95, 95% CI 2.6 to 8.8) and delivery at a tertiary referral centre (aOR 1.35, 95% CI 0.3 to 5.0).

**Conclusion:** We observed high neonatal mortality in both urban and rural West African settings, even among women considered to be at low risk of complications. Intrapartum related asphyxia, congenital malformations and low birth weight were important contributors to early neonatal deaths. There is an urgent need for targeted interventions to address preventable risk factors linked to neonatal mortality in West Africa, especially intrapartum related asphyxia.

## Introduction

The neonatal period, which spans the first 28 days of life, is the most vulnerable time in a child’s development (1,2). Although over the last three decades global neonatal mortality decreased by 50% (3), the proportion of global under-five deaths occurring during the neonatal period has increased from 40% in 1990 to 47% in 2021(4,5). In 2022, the global neonatal mortality was 17 per 1,000 live births, accounting for 2.3 million newborn deaths (6,7). From the limited data available, Sub-Saharan Africa (SSA) remains the region with the highest neonatal mortality, with 27 deaths per 1000 live births, representing an eleven-fold higher risk compared to high-income countries(8,9). West and Central Africa have the highest burden of neonatal mortality within SSA and globally, with 30 deaths per 1000 live births, and the slowest reduction rate of all global regions (10).

The United Nations (UN) Sustainable Development Goal (SDGs) target 3.2 aims to reduce neonatal mortality to less than 12 per 1,000 live births in every country by 2030. This goal highlights the critical importance of neonatal mortality not only as an indicator of child health and overall well-being, but also as a reflection of broader social and economic development. However, achieving this target requires focused efforts to address the most common causes of neonatal deaths (11), which globally include preterm birth, intrapartum related asphyxia, and neonatal infections (12). In SSA, these causes account for 28.5%, 22.5% and 18.4% of neonatal deaths, respectively (13). Although the exact distribution varies by setting, this pattern is generally consistent across SSA, which underscores the need to tailor preventative neonatal services (14–16)).

Neonatal mortality is influenced not only by direct medical causes but also by broader underlying factors at the family, community, and societal levels. These factors, often associated with low education and poverty, include limited maternal awareness about health seeking, poor maternal nutrition, inadequate water and sanitation facilities and insufficient access to basic health services (17,18). Maternal risk factors for neonatal deaths vary across populations but generally include maternal age, lack of antenatal care during pregnancy, antenatal infections, as well as complications during labour and delivery, including prolonged and assisted deliveries. Neonatal factors consistently associated with neonatal mortality include multiple gestation, prematurity, low birth weight (LBW), congenital malformations, low Apgar scores at birth (reflecting intrapartum related asphyxia), home delivery, sepsis, and hypothermia (19–23). Despite good understanding of generic risk factors, there are still evidence gaps for context-specific factors associated with neonatal mortality in West Africa, which is essential to inform maternity and neonatal health policies and programmes in this high-mortality burden region.

Our study aimed to describe the clinical epidemiology of neonatal mortality in The Gambia and Burkina Faso, focusing on the incidence, timing, and associated risk factors.

## Methods

### Study design

This study comprises of secondary data analyses from the PregnAnZI-2 trial (24), which was a phase III, double-blind, placebo-controlled randomised clinical trial conducted in The Gambia and Burkina Faso between October 2017 and May 2021 (clinicaltrials.gov ref:NCT03199547). Approximately 12,000 pregnant women in labour received either a single oral dose of azithromycin or placebo. The trial methods have been extensively described (24). The primary analysis found no difference in the primary composite endpoint of neonatal mortality and sepsis between the intervention and placebo arms (25). Therefore, in this secondary analysis, we have included all neonatal deaths regardless of trial arm or whether participants met PregnAnZI-2’s primary endpoint definition, which excluded deaths deemed unlikely to be influenced by intra-partum azithromycin [specifically intrapartum-related asphyxia, major congenital malformations and low birth weight]. By including all neonatal deaths, these analyses provide a comprehensive understanding of neonatal deaths in our cohort, which is broadly representative of the general population in The Gambia and Burkina Faso.

### Study setting

Participants were enrolled at ten government health facilities in The Gambia and Burkina Faso, West Africa. In The Gambia, two facilities located in the urban western region participated: Bundung Maternal and Child Health Hospital (a secondary care facility) and Serekunda Health Centre (a primary care facility). Bundung Maternal and Child Health Hospital provided emergency obstetric care, including caesarean sections, and only on rare occasions were women referred to the tertiary level Edward Francis Small Teaching Hospital, the national neonatal referral centre, 15.5 Km away. The hospital also offered WHO level 2 care for small and sick newborns, including oxygen, intravenous (IV) antibiotics, and fluids. Serekunda Health Centre, in contrast, provided only basic obstetric care and women requiring caesarean section were referred to Kanifing General Hospital, 3.4 Km away. Serekunda Health Centre provided essential newborn care and any unwell neonates were referred to either Kanifing General Hospital or Edward Francis Small Teaching Hospital, where WHO level 2+ care including oxygen, bubble CPAP, IV antibiotics and fluids were available. During the trial, neonates who became unwell post-discharge were admitted to either Bundung Maternal Child Health Hospital or to the Medical Research Council The Gambia (MRCG) Clinical Services Department, where WHO level 2 neonatal care was available (24).

In Burkina Faso, women were enrolled at eight peripheral health facilities located in the rural districts of Nanoro and Yako. All health facilities provided primary care, including basic obstetric care except for The Centre Medical Saint Camille de Nanoro (CMA), the referral hospital located <25Km away, where caesarean sections could be performed. Newborns requiring hospital admission were transferred to CMA, which offered WHO level 2+ small and sick newborn care (26). All neonatal care followed local clinical guidelines and was delivered at the discretion of health facility personnel. Newborns who became unwell post-discharge and returned to the trial facilities were also referred to CMA and transported by the trial team.

The Neonatal Mortality Rate (NMR) in 2020 in both countries was similarly high at 25.5 deaths per 1000 live births in Burkina Faso and 25.7 deaths per 1000 live births in The Gambia (27). The climate of both countries is typical of the sub-Sahel region, with a long dry season from November to May and a short rainy season between June and October. The population of the catchment area in The Gambia is representative of the main ethnic groups; Mandinka, Wolof and Fula, with high illiteracy rates, up to 55% among women (24). In Burkina Faso, the trial site is predominantly populated by the Mossi ethnic group (90%), with mostly subsistence farmers and cattle-farmers (about 50%) and more than 75% illiteracy rate (28).

### Participants

Participants included all pregnant women aged >16 years who were enrolled in the PregnAnZI-2 clinical trial and delivered at a study site either in The Gambia or Burkina Faso. Maternal exclusion criteria at the time of screening and enrolment included: planned (elective) caesarean section; antenatally detected major congenital malformation; known serious acute or chronic maternal morbidity (e.g., HIV infection); known macrolide allergy or maternal use of drugs known to prolong QT interval during the preceding two weeks (e.g., erythromycin, chloroquine). (24) All live-born offspring were included, with exclusion of stillborn infants who were identified by health facility staff following manual ascertainment of an absent heart rate and breathing at delivery.

### Study procedures

Written consent to participate in the study was taken during antenatal visits and verbally confirmed during labour. Socio-demographic, antenatal and intrapartum clinical data were collected as per study specific procedures. Apgar scores were documented prospectively at 1 minute and 5 minutes by health-facility staff following standard care, including resuscitation as per the Helping Babies Breathe protocol. Birth weight was measured by trained research nurses as soon as possible after delivery and within 24h using a calibrated digital scale to 0.1 kg precision. A thorough clinical assessment of the newborn, including examination for congenital malformations, was carried out by a research clinician or paediatrician 4–24h after delivery and prior to discharge or transfer. Newborns requiring additional care were admitted to the neonatal ward if available, or referred to a higher-level neonatal care facility, at the discretion of the health-facility clinician.

Women and neonates were followed up passively to 28 postnatal days with active surveillance at day 28 (+/-2 days) either at home (in The Gambia) or the nearest health facility study site (in Burkina Faso). During the follow-up period women were encouraged to contact the research team with any maternal or newborn health concerns. They were provided with mobile phone credit to encourage communication and travel expenses.(24)

### Outcomes and variables of interest

The primary outcome of interest was neonatal mortality (binary variable of yes or no), defined as any death of a live born neonate which occurred within 28 days after delivery. Neonatal deaths were further classified as early neonatal deaths (occurring within 72h after birth), and late neonatal deaths (occurring between 72h and day 28).

A low Apgar score (defined as <7 out of 10) at either 1 or 5 minutes after birth was taken as a proxy for intra-partum related asphyxia (Beloum et al, 2025, in press). Apgar score is a clinical score consisting of prospective assessment of breathing, heart rate, colour, tone and response to tactile stimuli.

For this study, “congenital malformations” were defined as structural anomalies that were easily recognisable on physical examination. The contemporaneous descriptions by research clinicians were categorised and coded as per the ICD-11 classification system (Graves S et al, 2025 – in press) by an experienced neonatologist. Minor, major and multiple malformations were represented by a binary variable (malformation or no malformation).

### Data management and statistical analysis

Data were collected electronically using an electronic data capture system (REDCap) and hand-held mobile devices. All data were pseudo-anonymised with internal consistency checks and validated according to source documents.

Data were analysed using R version 4.4.2 (2024-10-31 ucrt). The neonatal mortality rate was calculated as the number of neonatal deaths per 1,000 live births for the full cohort, with further stratification by country. To visualise timing of neonatal deaths, we used Kaplan-Meier survival curves, and survival probabilities were compared between countries using the log-rank test, providing a clear depiction of survival probabilities over the study period.

We used logistic regression to explore associations between independent “risk factors” and early neonatal mortality. Only early deaths were considered, (i.e., only deaths within 72hrs), as aetiology of risk factors are different for early versus late neonatal deaths and there were too few late neonatal deaths to analyse this outcome. Risk factors were selected for inclusion in regression models according to both their availability in the PregnAnZI-2 trial dataset and their relevance for early neonatal mortality, as reported in the literature. The baseline category of categorical variables was chosen to reflect either a “normal pregnancy and delivery” (e.g., normal birth weight, non-extremes of parity) or the most prevalent grouping (e.g., ethnicity). As country and ethnicity are co-linear, only ethnicity was included in adjusted models to provide more granular insights. To avoid over adjustment for variables on the same causal pathway, variables with *p* <0.2 on univariate analysis were included in adjusted regression models only if they occurred before or at the same timepoint as the independent variable of interest (29). Results for the unadjusted and adjusted models were expressed as crude and adjusted odds ratios (OR), respectively, with 95% confidence intervals (CI) and p-values. Statistical significance for variables associated with mortality on the adjusted regression model was defined as *p*<0.05. Missing data were < 1% for all independent variables and available case analysis was used.

## Results

### Overview of participants and neonatal mortality

In total, 11,980 pregnant women gave birth to 12,105 live-born neonates, of whom 1.2% (144/12,105) died during the neonatal period (Figure 1). 59% (85/144) of the deaths occurred in The Gambia and 41% (59/144) in Burkina Faso. The neonatal mortality rate (NMR) for the whole cohort was 11.5 per 1,000 live births (144/12,105), 12.5 per 1000 live births (85/6,773) in The Gambia and 11.1 per 1000 live births (59/5,332) in Burkina Faso (Figure 1). 82% (118/144) of all neonatal deaths occurred during the early neonatal period (Figure 2), 79% (67/85) in The Gambia; and 86% (51/59) in Burkina Faso.

**Figure 1.**
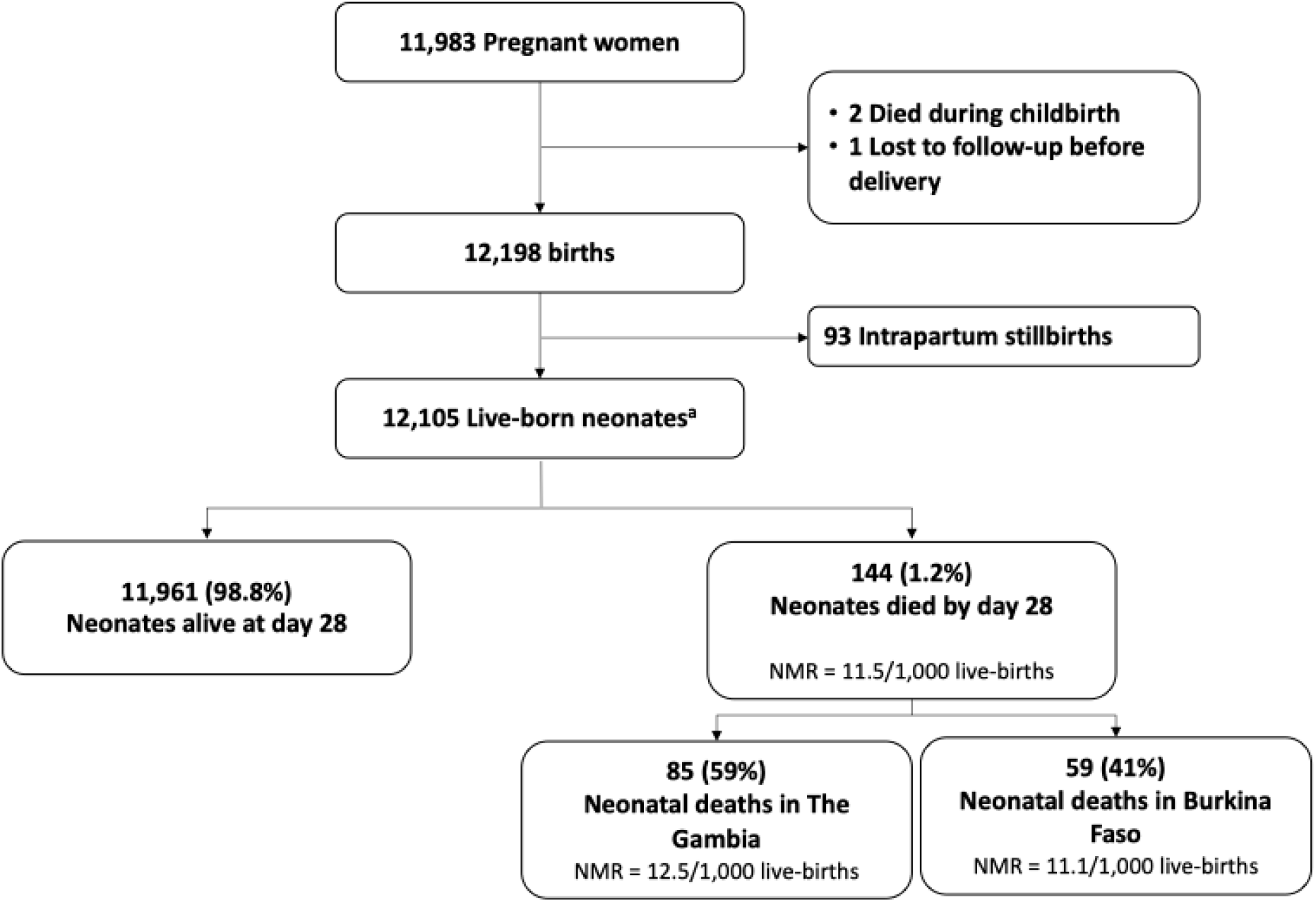
Overview of population and neonatal mortality from a West African cohort of pregnant women considered to be at low risk of complications at onset of labour NMR = Neonatal mortality rate a) 55.2% (6,688/12,105) of live-born neonates were in The Gambia and 43.6% (5,273/12,105) in Burkina Faso

**Figure 2.**
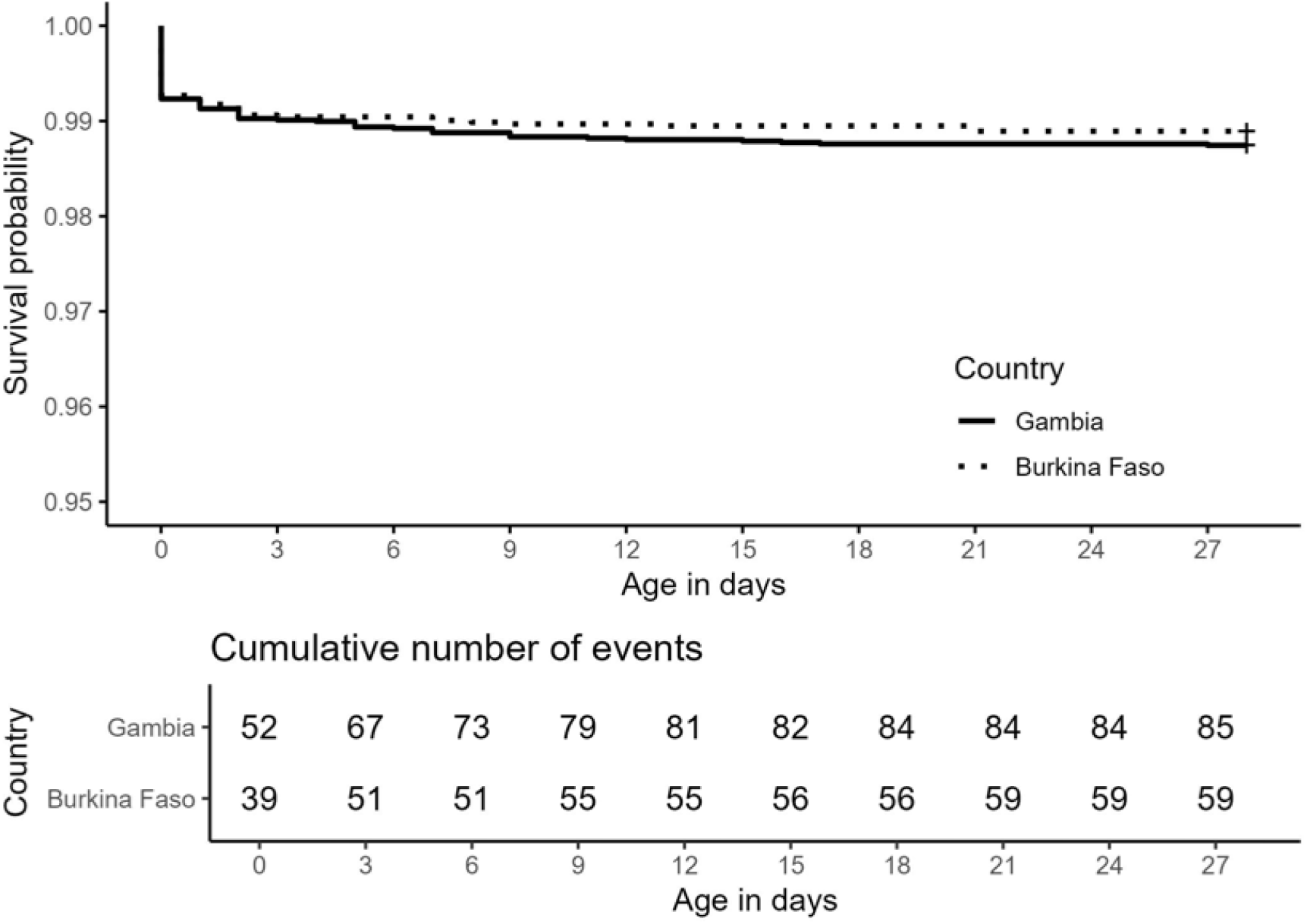
Kaplan Meir graph for whole cohort and stratified by country

### Factors associated with early neonatal mortality

A history of stillbirth in a previous pregnancy was the only maternal or antenatal factor associated with early neonatal mortality (cOR 4.95, 95% CI 2.6-8.8, *p*<0.001). A low Apgar score at 1 minute (aOR 49.85, 95% CI (32.2-77.6, *p* <0.001) and 5 minutes (aOR 92.38, 95% CI 56.6-151.3, *p*<0.001) greatly increased the odds for early neonatal mortality. Neonates with a congenital malformation had 11-fold increased odds of early neonatal death compared to those without a malformation (aOR 11.44, 95% CI 6.4-19.4, *p*<0.001). Birth weight was also associated with early neonatal mortality (*p<*0.001) with four-fold increased odds for LBW neonates (aOR 4.09, 95% CI 2.6-6.3, *p*<0.001) (Table 1).

**Table 1.**
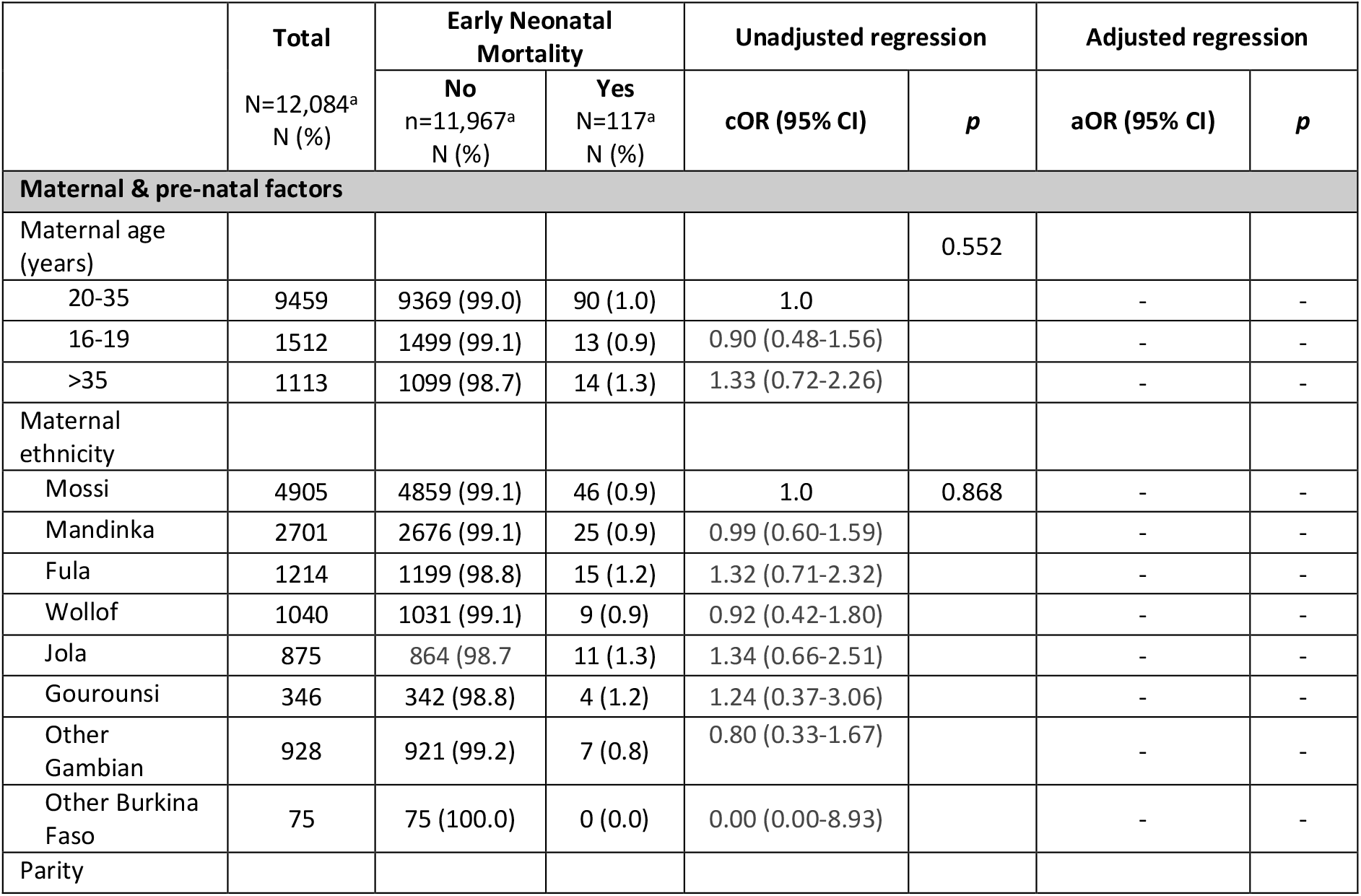
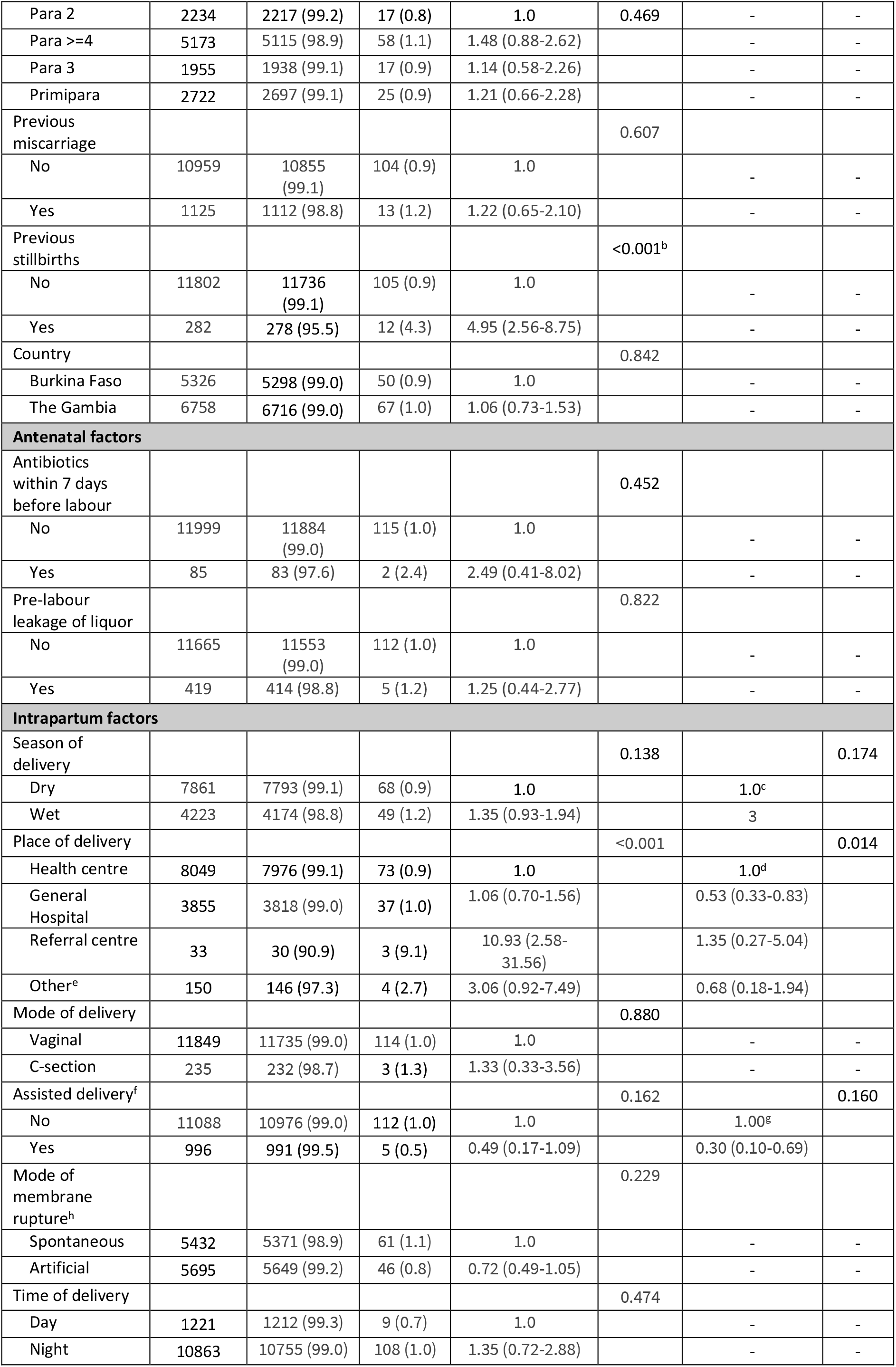
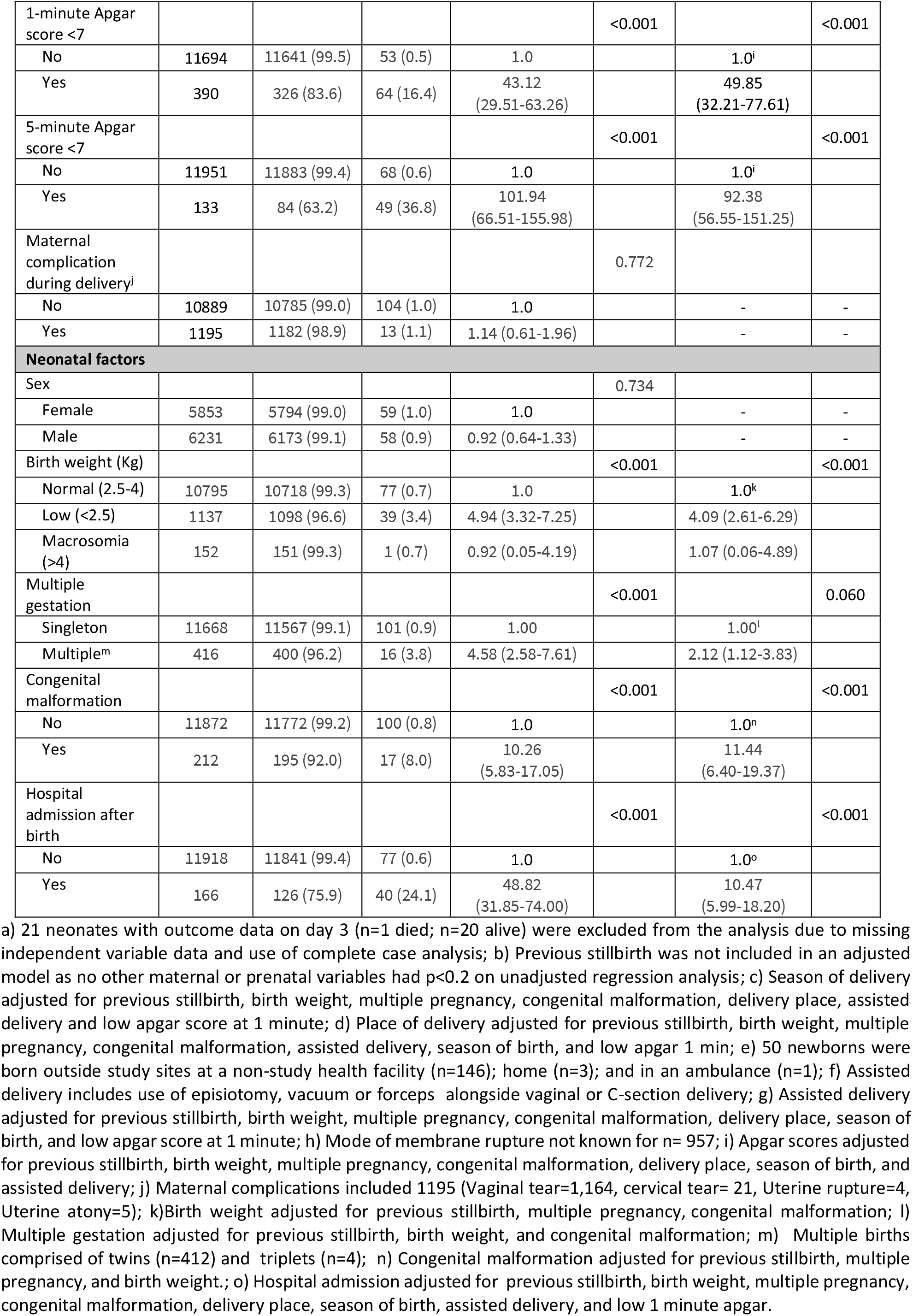
Maternal and neonatal socio-demographic and clinical factors associated with early neonatal mortality (0-72h) following health-facility delivery in The Gambia and Burkina Faso.

## Discussion

This multi-country study provides important insights into neonatal mortality among facility-born neonates in two West African countries. We examined a large cohort of pregnant women classified as healthy during labour, hence our findings reflect outcomes from low-risk pregnancies. Despite this selected population, the observed NMR was relatively high, especially during the early neonatal period, which underlines persistent challenges in maternity and neonatal care in the region. We identified several risk factors associated with early neonatal deaths, many of which are commonly identifiable during pregnancy, such as a history of stillbirth, LBW representing growth restriction or prematurity, and presence of a congenital malformation. Low Apgar scores (1-minute and 5-minute) showed strong associations with early neonatal mortality, suggesting that intrapartum-related asphyxia is a leading contributor to neonatal deaths in The Gambia and Burkina Faso.

In our cohort, the overall NMR was 11.5 per 1,000 live births, significantly lower than national estimates for both The Gambia (25.7 deaths per 1000 live births) and Burkina Faso (25.5 deaths per 1000 live births)(27). This relatively lower rate was expected as women with known acute or chronic pregnancy complications were excluded (25). In addition, women and neonates in our cohort likely benefited from participating in a well-run clinical trial with enhanced access to antenatal, intrapartum and postnatal care provided by the research team (“trial effect”) (30). The NMR reported in our study is comparable to rates reported in middle-income countries such as Indonesia (NMR 11.5), Morocco (NMR 11.4), and South Africa (NMR 11.2)(31), supporting the evidence that improved maternal health and higher quality perinatal care can substantially reduce neonatal mortality in LMICs (32,33).

More than four out of five neonatal deaths in our cohort occurred within the first 72 hours after birth,, consistent with findings from population-based studies in LMIC (34,35). More specifically, a prospective global network study conducted across five low-income countries (Kenya, Zambia, Guatemala, India, Pakistan) and one middle-income country (Argentina), reported that over half of neonatal deaths occurred within the first 24 hours of life (36). This timing underscores the urgent need for high-quality small and sick newborn care starting immediately after birth(37). This pattern of neonatal mortality occurring early in the neonatal period has also been observed in settings with advanced healthcare systems (38), despite their significantly lower mortality rates, thus underscoring the importance of integrating high-quality antenatal and intrapartum care strategies to optimise foetal health and improve early neonatal outcomes.

Evidence of risk factors for early neonatal mortality in West Africa is largely limited to demographic surveys, with limited data from large, prospective cohorts such as ours. We identified that low 1-minute and 5-minute Apgar scores are strong risk factors for early neonatal mortality, highlighting the contribution of intrapartum-related asphyxia towards early neonatal deaths in The Gambia and Burkina Faso. This finding is consistent with Gambian data from over a decade ago, which attributed 31% of neonatal deaths to intrapartum-related asphyxia (39). A linked analysis also using PregnAnZI-2 trial data further supported this association, showing that 40% of neonatal deaths in The Gambia were associated with intrapartum-related asphyxia, reinforcing its central role in the neonatal mortality pathway (Beloum et al, in press). However, due to the nature of our study and data limitations, we were unable to assess the severity of intrapartum-related asphyxia, the quality of neonatal resuscitation provided, or the presence of infective co-morbidities. Currently there are no proven interventions in African settings to improve neonatal survival or long-term neuro-developmental outcomes following intrapartum-related asphyxia(40). Our findings, therefore, underscore the urgent public health need for effective intrapartum monitoring, timely and safe delivery, and provision of high quality neonatal resuscitation (41,42) as core strategies to reduce early neonatal mortality in West Africa.

We identified several other neonatal risk factors which, if detected early in pregnancy, could enable closer antenatal monitoring, optimisation of antenatal management and planning for safe delivery, thus reducing the risk of intrapartum related asphyxia and early neonatal deaths. Newborns with an easily identifiable congenital malformation had 11-fold increased odds of early mortality in our cohort. Although our data was limited to easily recognisable malformations, and concealed anomalies may have been missed, our finding is consistent with that of a similar Gambian study conducted in 2013-2014 which identified 36-fold increased risk of neonatal mortality for newborns with major malformations (43). The higher mortality-risk in this earlier study probably reflects differences in the severity of malformations, as our analysis included both minor and major anomalies. Musculo-skeletal malformations were the most common type of malformation in our cohort (Graves et al, in press), which are known to have higher risk of breech presentation (44) and subsequent asphyxia (45). Regardless of the mechanisms, early antenatal detection and optimised intrapartum management of pregnancies affected by malformations is critical to mitigate the risk of early neonatal mortality (46– 48). LBW due to prematurity, growth restriction or both, is also a major contributor to neonatal mortality in all settings, as identified by the recent Lancet Small Vulnerable Newborn (SVN) series (49). Our finding of a four-fold increased odds of early mortality for LBW neonates is consistent with this. However, we cannot comment on the SVN phenotype in our cohort as antenatal and postnatal gestational age estimates were not available in our dataset.

In addition to the limitations highlighted above, we acknowledge the potential bias introduced by half of our cohort receiving intrapartum azithromycin. Although neonatal mortality rates were similar between trial arms, we were unable to assess potential differences in the causes, timing of deaths or risk factors for death associated with azithromycin use. Although our cohort included women from urban and rural West African settings, we cannot comment on potential intra-country variations in neonatal mortality. For example, a recent HDSS-based Gambian study identified rural residence as a risk factor for neonatal mortality in the community (50) and our study included Gambian urban health facilities and Burkinabe rural health facilities. Finally, the risk factors analysis was constrained by the variables available in the study. Important factors such as detailed socio-economic data, antenatal care and postnatal practices (e.g., feeding methods, kangaroo care) were not available, limiting our ability to detect risk or protective factors for survival.

## Conclusion

Early neonatal mortality remains unacceptably high in both urban and rural West African settings, even among pregnant women considered to be at low risk of complications. Intrapartum-related asphyxia is the leading contributor to early neonatal deaths, with congenital malformations and LBW also playing significant roles. To achieve the SDG target 3.2 within the next five years in both countries, targeted, evidence-based interventions during pregnancy and labour should be urgently implemented, particularly those aimed at preventing and managing intrapartum related asphyxia.

## Data Availability

Data may be obtained from a third party and are not publicly available. The clinical data has been collected following provision of informed consent under the prerequisite of strict participant confidentiality. Qualified researchers may request access with the Gambia Government MRC Joint Ethics Committee. The review process and release of data will be facilitated by MRC Unit The Gambia (http://www.mrc.gm/) through the Head of Governance at MRCG. Access will not be unduly restricted.

## DECLARATIONS

### Ethical approval

The parent trial was approved by The Gambia Government/MRCG (Medical Research Council Unit The Gambia) Joint Ethics Committee, the Comité de Ethique pour la Recherche en Santé (CERS), the Ministry of Health of Burkina Faso, and the LSHTM Ethics Committee. All women provided written informed consent during ante-natal care visits and were free to withdraw at any time.

### Authors’ contributions

AR conceptualised this study with input from UN, HB, UdA and RT. Data collection was co-ordinated by BC, JDB and the PregnAnZI-2 field teams in The Gambia and Burkina Faso. TR performed the analysis with full access to the data. UN drafted the initial manuscript with input from HB and AR. All authors contributed to the final version. AR gave oversight to the work as guarantor and accepts full responsibility for finished work and controlled the decision to publish.

### Data sharing statement

Data may be obtained from a third party and are not publicly available. The clinical data has been collected following provision of informed consent under the prerequisite of strict participant confidentiality. Qualified researchers may request access with the Gambia Government/MRC Joint Ethics Committee. The review process and release of data will be facilitated by MRC Unit The Gambia (http://www.mrc.gm/) through the Head of Governance at MRCG. Access will not be unduly restricted.

## Acknowledgements

We acknowledge the project managers and all the field, data-management, and research support teams at MRCG and CRUN for providing support to conduct the PregnAnZI-2 trial. We also thank the leadership boards and staff at the study sites for their support. We are grateful to all the mothers and their neonates who participated in this study.

